# Knowledge and Perceptions of Hypertension Among Patients in a Cardiology Clinic

**DOI:** 10.1101/2025.08.25.25334408

**Authors:** Fatima Khurshid, Muhammad Owais, Iqra Tahir Dar, Fiza Ameer, FNU Absar, Ayesha Khurshid, Raheel Ahmed, Bravein Amalakuhan

## Abstract

**Background:** Hypertension is a leading global health concern, contributing significantly to cardiovascular morbidity and mortality. Despite its serious complications, many patients have insufficient knowledge of the condition, its management, and associated risks.

**Objective:** This study aimed to assess the knowledge and perceptions of hypertension among patients attending a cardiology clinic, highlighting misconceptions and knowledge gaps that may influence treatment adherence and disease outcomes.

**Methodology:** A cross-sectional study was conducted at the cardiology clinic of Liaquat University Hospital, Pakistan, involving 200 patients selected through convenience sampling. Data were collected using the validated Hypertension Knowledge-Level Scale (HKLS), which includes six sub-dimensions: definition, medical treatment, medication compliance, lifestyle, nutrition, and complications. Normality of the data was tested with the Shapiro-Wilk test. As the data were not normally distributed, descriptive statistics (frequencies and percentages) were analyzed using SPSS.

**Results:** Of the 200 participants (mean age: 41 years), the overall mean HKLS score was 14.1 out of 22, indicating moderate knowledge. While 61–64.5% correctly identified the definition of hypertension, misconceptions persisted. Notably, 40% incorrectly believed medication is required only during symptomatic episodes, 67% thought hypertension in older adults does not require treatment, and 79% considered frying a healthy cooking method. Awareness of complications varied, with 70.5% recognizing heart attack risk but only 49% identifying stroke risk.

**Conclusion:** The findings demonstrate significant knowledge gaps and misconceptions regarding hypertension. Targeted educational interventions are essential to improve treatment adherence, encourage lifestyle modification, and inform healthcare policy for better patient outcomes.

## INTRODUCTION

Hypertension is characterized by a consistent blood pressure measurement of 140/90 mmHg or above. It is a major contributor to early mortality on a global scale, with a goal to decrease its occurrence by 33% from 2010 to 2030. The issue is especially difficult because it typically shows no symptoms until it reaches a point where it can lead to serious problems like stroke, heart attack, and kidney failure [1]. Elevated systolic blood pressure, which damages coronary arteries and increases the risk of myocardial infarction, is an important predictor of cardiovascular disease risk. It increases the burden on the heart over time, causing dilatation and weakening of the left ventricle. As blood pressure rises, the probability of a serious cardiovascular incident increases [2]. Hypertension is a worldwide public health hazard connected to urbanization and socioeconomic factors that encourage sedentary lifestyles. Controlling hypertension and avoiding consequences is difficult owing to a lack of awareness and poor drug adherence. Patient education is critical in controlling hypertension, and initiatives are underway to raise public knowledge of the dangers of uncontrolled blood pressure. Medical personnel must comprehend the significance of systolic blood pressure in hypertension prevention and management [3]. Thus, assessing a patient’s comprehension and identification of hypertension is critical. The World Health Organization calculates that 1.28 billion adults between the ages of 30 and 79 worldwide have high blood pressure, with most living in low- and middle-income nations. Although it is common, hypertension frequently goes undetected and untreated, resulting in a significant impact on health [1]. Recent research has emphasized the significance of patient understanding, beliefs, and behaviors (KAP) regarding hypertension. Sa’adeh and colleagues (2018) examined hypertensive patients’ knowledge, attitudes, and practices regarding the prevention and early detection of chronic kidney disease in Palestine. Higher levels of knowledge and attitude were linked to improved adherence to preventive measures, as indicated by the study findings [4]. The study examined community pharmacists in China to assess their knowledge, attitude, and practice towards patient education on hypertension, highlighting a positive attitude alongside insufficient knowledge and practice Chen et al., 2022) [5]. A WHO study in 2018 offered a detailed look at how to manage hypertension and highlighted the significance of educating patients to enhance treatment results. The study examines patient attitudes and perceptions of hypertension in three low and middle-income countries. It found that patients prefer Home Blood Pressure Monitoring (HBPM) and a medication self-titration plan to address cost and safety. However, the study identified knowledge gaps in hypertension causes, measurement, and treatment adherence. It suggests targeted interventions and education programs to bridge these gaps and improve blood pressure management. The study also compares knowledge and perception among patients with different demographic characteristics [6]. A systematic review of studies reveals barriers to hypertension awareness, treatment, and follow-up, including lack of resources, time, knowledge gaps, motivation, and beliefs about treatment consequences. These barriers differ between high-income and low-and middle-income countries. The study suggests targeted interventions and methodologically rigorous studies to improve hypertension control (Khatib et al, 2014) [7]. This study aims to fill knowledge gaps in patients about hypertension, identifying misunderstandings and potential risks. It evaluates patients’ understanding of hypertension, its causes, symptoms, risk factors, and treatments. The research also investigates how patients perceive and feel about hypertension, identifying potential risks and misunderstandings. It also examines the impact of understanding on self-care actions and demographic features. The study aims to improve patient education and healthcare policies.

While the importance of patient knowledge is established globally, specific misconceptions prevalent within the local Pakistani context remain underexplored. Therefore, this study aims to answer the following question: What is the level of knowledge regarding hypertension definition, treatment, medication compliance, lifestyle, diet, and complications among patients attending a tertiary care cardiology clinic in Pakistan?

## MATERIALS AND METHODOLOGY

This research employed a cross-sectional study design to assess hypertension knowledge among patients attending the cardiology clinic of Liaquat University Hospital, a tertiary care facility in Hyderabad/Jamshoro, Pakistan, which serves a diverse urban and rural population.

### Sampling and Participants

Participants were selected through a non-probability convenience sampling method. The study included adult patients (aged 18 and over) with or without a prior diagnosis of hypertension who were present at the clinic and consented to participate. Data collection took place over a one-month period, from May to June 2024. The sample size of 200 was determined based on the patient flow during the study period and feasibility.

### Data Collection Instrument

Data were collected using the Hypertension Knowledge-Level Scale (HKLS), developed and validated by Erkoc et al. (2012) in Turkey [8]. The original scale demonstrated high reliability, with a Cronbach’s alpha coefficient of 0.83. For this study, the questionnaire was translated from English into the local languages (Urdu and Sindhi) by two bilingual experts and then back-translated into English by a third independent expert to ensure linguistic and conceptual equivalence. The translated questionnaire was pilot-tested on 20 patients from a different clinic to check for clarity and comprehension, and minor wording adjustments were made. The internal consistency of the translated scale in our sample was found to be acceptable, with a Cronbach’s alpha of 0.76.

The HKLS consists of 22 items assessing six dimensions of hypertension knowledge: definition (2 items), medical treatment (4 items), medication compliance (4 items), lifestyle (5 items), nutrition (2 items), and complications (5 items). Responses are “Correct,” “Incorrect,” or “Don’t Know.”

### Data Collection Procedure

A team of four final-year medical students was trained as data collectors. The training involved a two-hour session covering the study’s objectives, ethical principles, obtaining informed consent, and a standardized, neutral method for administering the HKLS to avoid influencing participant responses. Data were collected via face-to-face interviews conducted in a semi-private area of the clinic’s waiting room to ensure confidentiality while patients were waiting for their appointments. This method was chosen to include participants with varying literacy levels.

## Data Analysis

Data were analyzed using SPSS version 26. The normality of the total HKLS score was evaluated using the Shapiro-Wilk test. The test indicated that the data were not normally distributed (p < 0.05). Therefore, descriptive statistics, including frequencies and percentages, were used to summarize participant characteristics and responses to each HKLS item. The total knowledge score was calculated for each participant (1 point for each correct answer, max score of 22).

## Ethical Considerations

The study protocol was approved by the LUMHS Research Ethical Committee (LUMHS/REC/-312). Written informed consent was obtained from all participants after explaining the study’s purpose. Participation was voluntary, and confidentiality was maintained by anonymizing all data.

## RESULTS

A total of 200 participants were included in the study, reflecting a diverse range of sociodemographic characteristics, with an emphasis on their hypertension management knowledge. The age distribution of the participants varied, with a significant portion falling in the 40-49 age group (41%), followed by those in the 20-29 age group (22%), 30-39 years group (16.5%), 50-59 years group (17.5%), and the smallest percentage in the 60-65 years group (3%), with an average age of 41 years. The participants’ residential origins varied, with 58.5% living in cities and 41.5% in rural regions. This blend emphasizes the range of environmental and lifestyle circumstances that may have an influence on their understanding and management of hypertension.

In terms of educational achievement, the cohort had a greater proportion of moderately to highly educated people: 36% had graduated, and 30% had completed upper secondary education. However, only a small proportion have elementary (7%) or secondary (4%) education, indicating a typically well-educated group. The gender distribution was nearly even, females (51.5%) over males (48.5). This near-even split minimizes gender-specific biases and verifies the study findings’ general application to all genders. Marital status found that the majority of participants (72.5%) were married, with unmarried persons (27.5%). A large proportion of the sample (74%) had a history of hypertension, whereas 26% had no prior diagnosis. A family history of hypertension was common, with 63.5% suggesting a relation to the disease and 36.5% reporting no such history. The occupation status was evenly split, with 49.5% unemployed and 50.5% employed. These sociodemographic data (Table 1) provide a comprehensive picture of the research population.

**Table 1:**
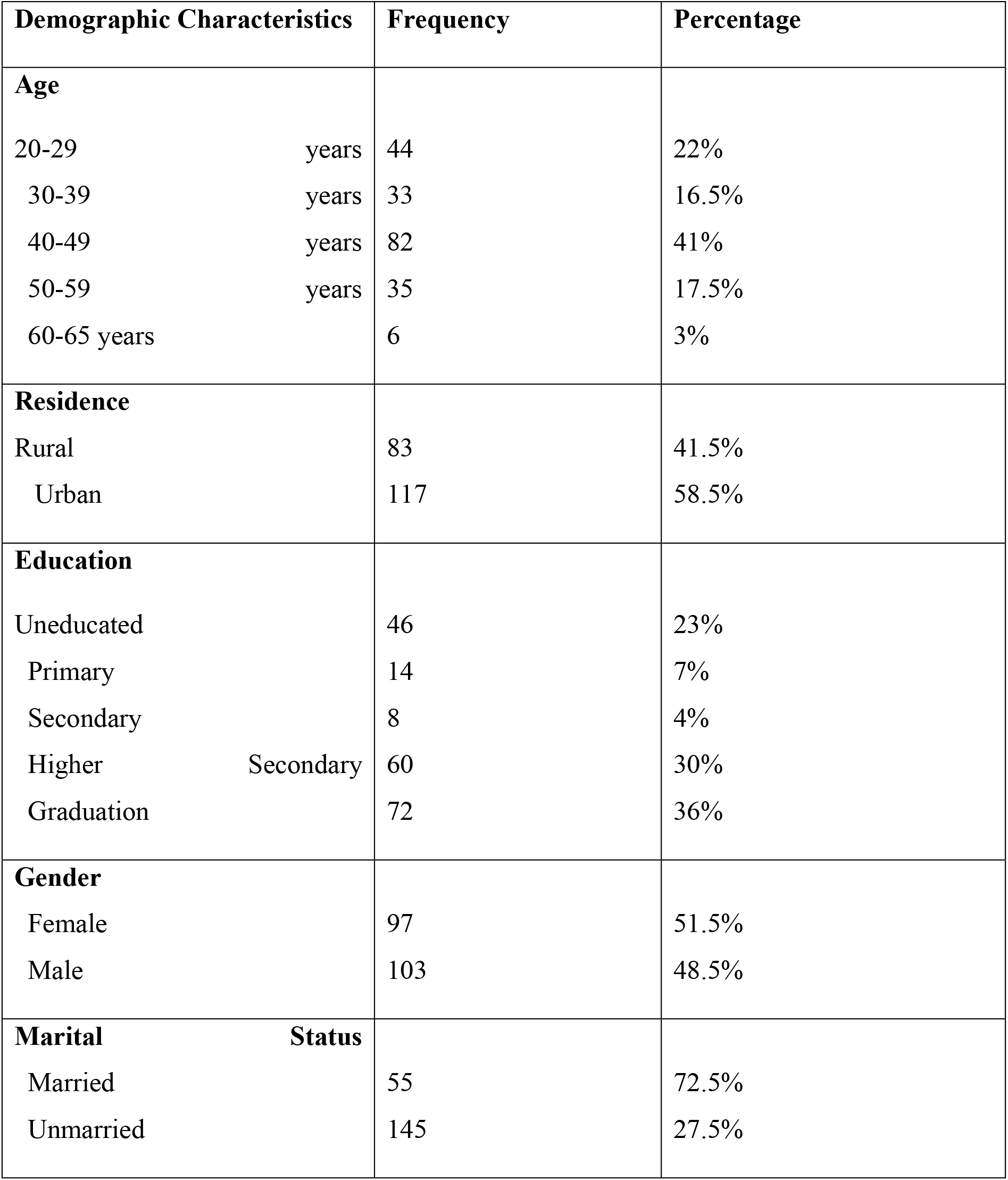
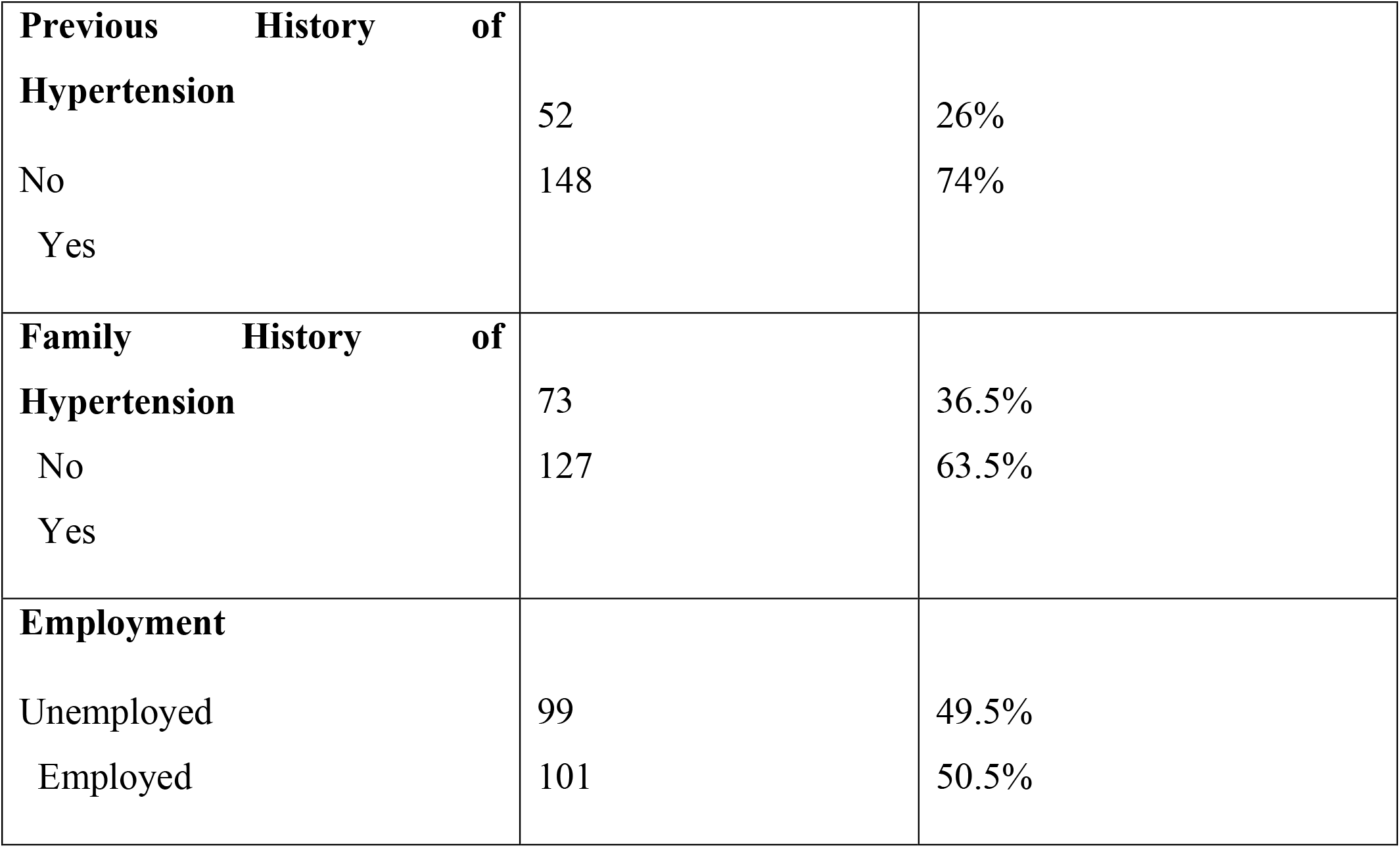
Sociodemographic characteristics of the participants. (N=200)

| Demographic Characteristics |  | Frequency | Percentage |
| --- | --- | --- | --- |
| **Age** |  |  |  |
| 20-29 | years | 44 | 22% |
| 30-39 | years | 33 | 16.5% |
| 40-49 | years | 82 | 41% |
| 50-59 | years | 35 | 17.5% |
| 60-65 years |  | 6 | 3% |
| **Residence** |  |  |  |
| Rural |  | 83 | 41.5% |
| Urban |  | 117 | 58.5% |
| **Education** |  |  |  |
| Uneducated |  | 46 | 23% |
| Primary |  | 14 | 7% |
| Secondary |  | 8 | 4% |
| Higher | Secondary | 60 | 30% |
| Graduation |  | 72 | 36% |
| **Gender** |  |  |  |
| Female |  | 97 | 51.5% |
| Male |  | 103 | 48.5% |
| **Marital Status** |  |  |  |
| Married |  | 55 | 72.5% |
| Unmarried |  | 145 | 27.5% |

|  |  |  |
| --- | --- | --- |
| **Previous History of Hypertension** |  |  |
| No | 52 | 26% |
| Yes | 148 | 74% |
| **Family History of Hypertension** |  |  |
| No | 73 | 36.5% |
| Yes | 127 | 63.5% |
| **Employment** |  |  |
| Unemployed | 99 | 49.5% |
| Employed | 101 | 50.5% |

Analysis of the Hypertension Knowledge-Level Scale (HKLS) responses (Table 2) revealed specific areas of strength and weakness in patient knowledge. For the Definition sub-dimension, 61% correctly identified high systolic pressure and 64.5% identified high diastolic pressure as indicators of hypertension. In the Medical Treatment sub-dimension, a high proportion (83.5%) agreed that medication should be taken as believed best, but there was a significant split on whether medication should be taken daily (44.5% correct) or only when feeling unwell (40% correct).

**Table 2:**
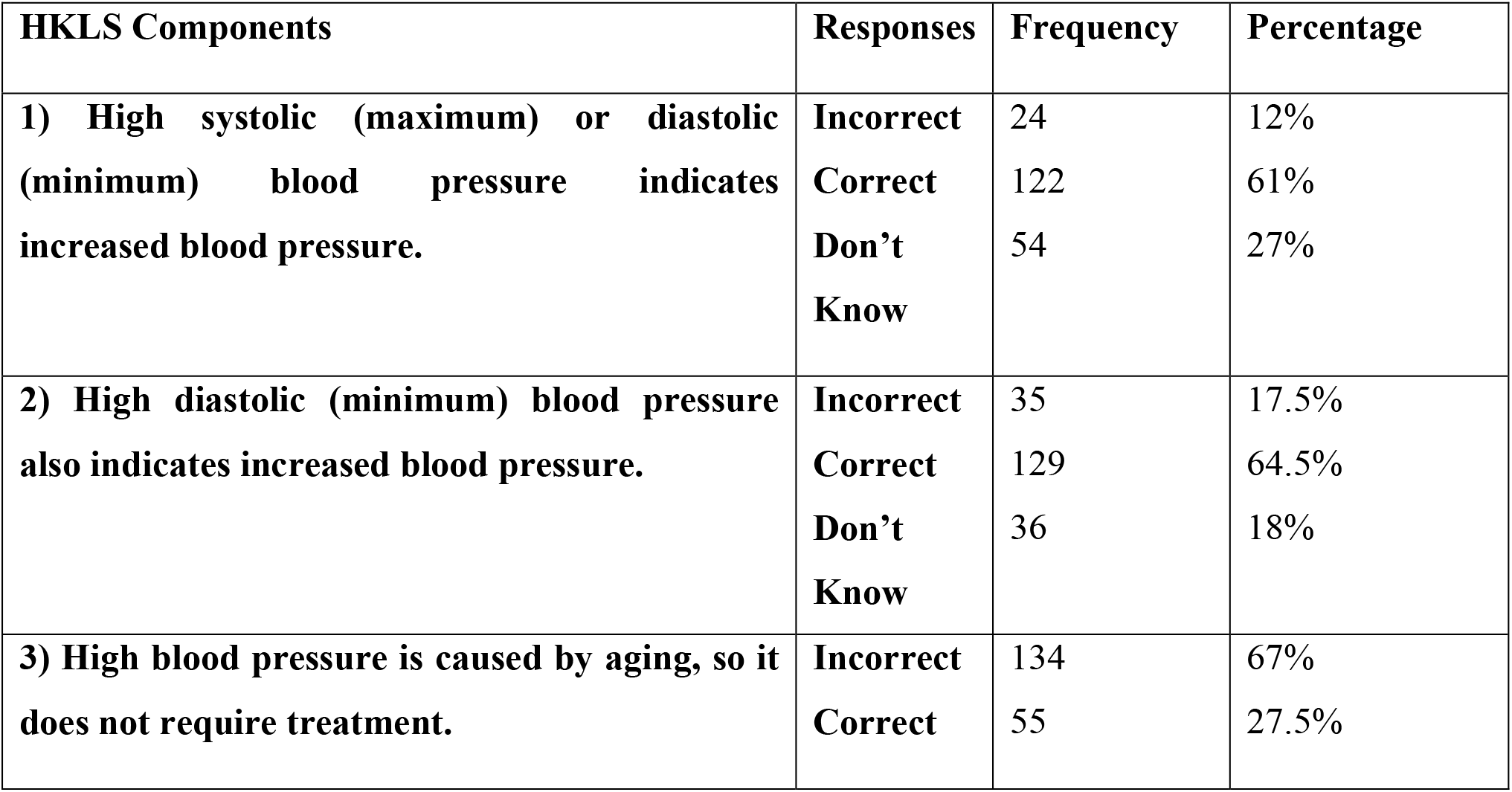
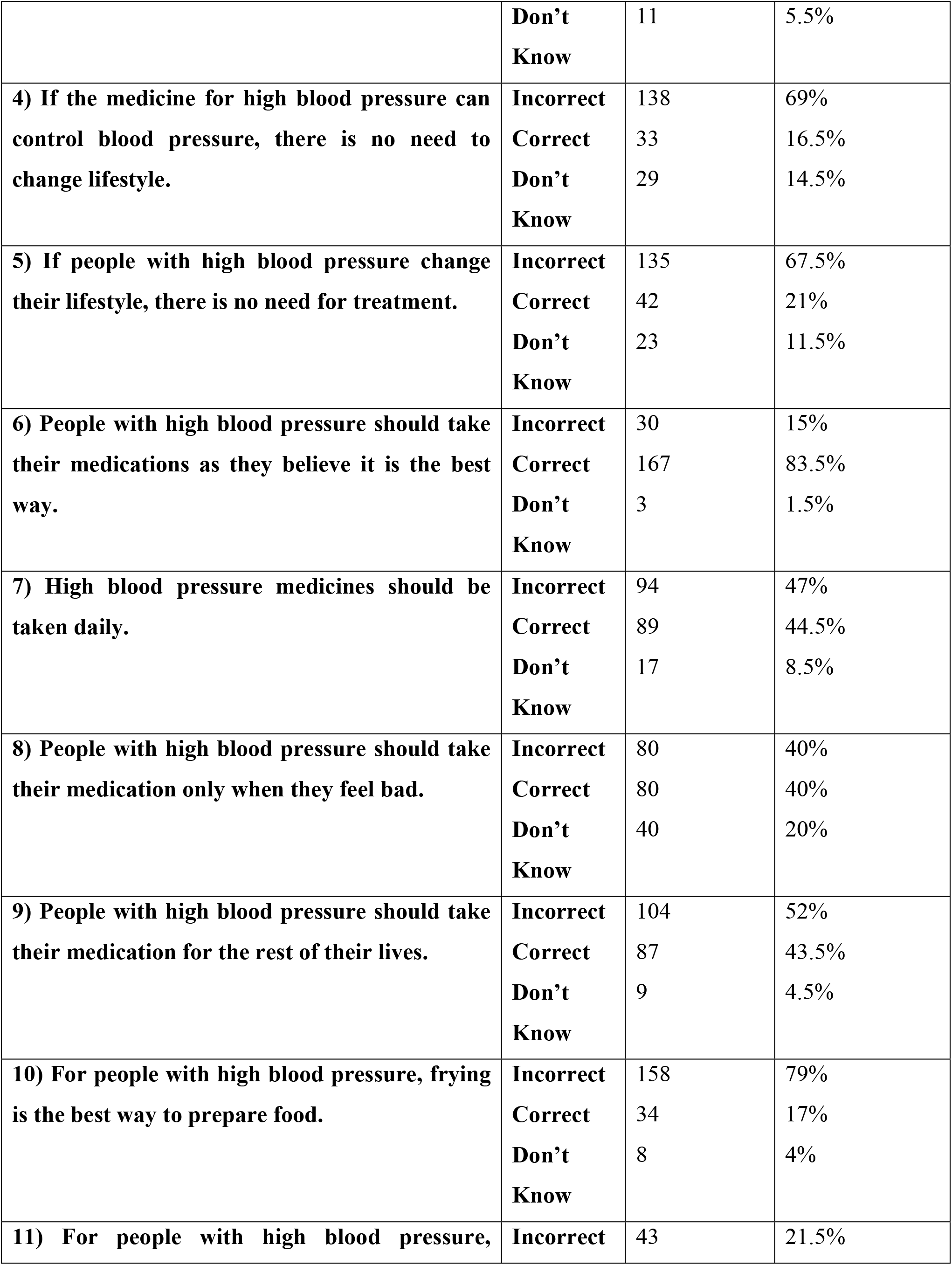
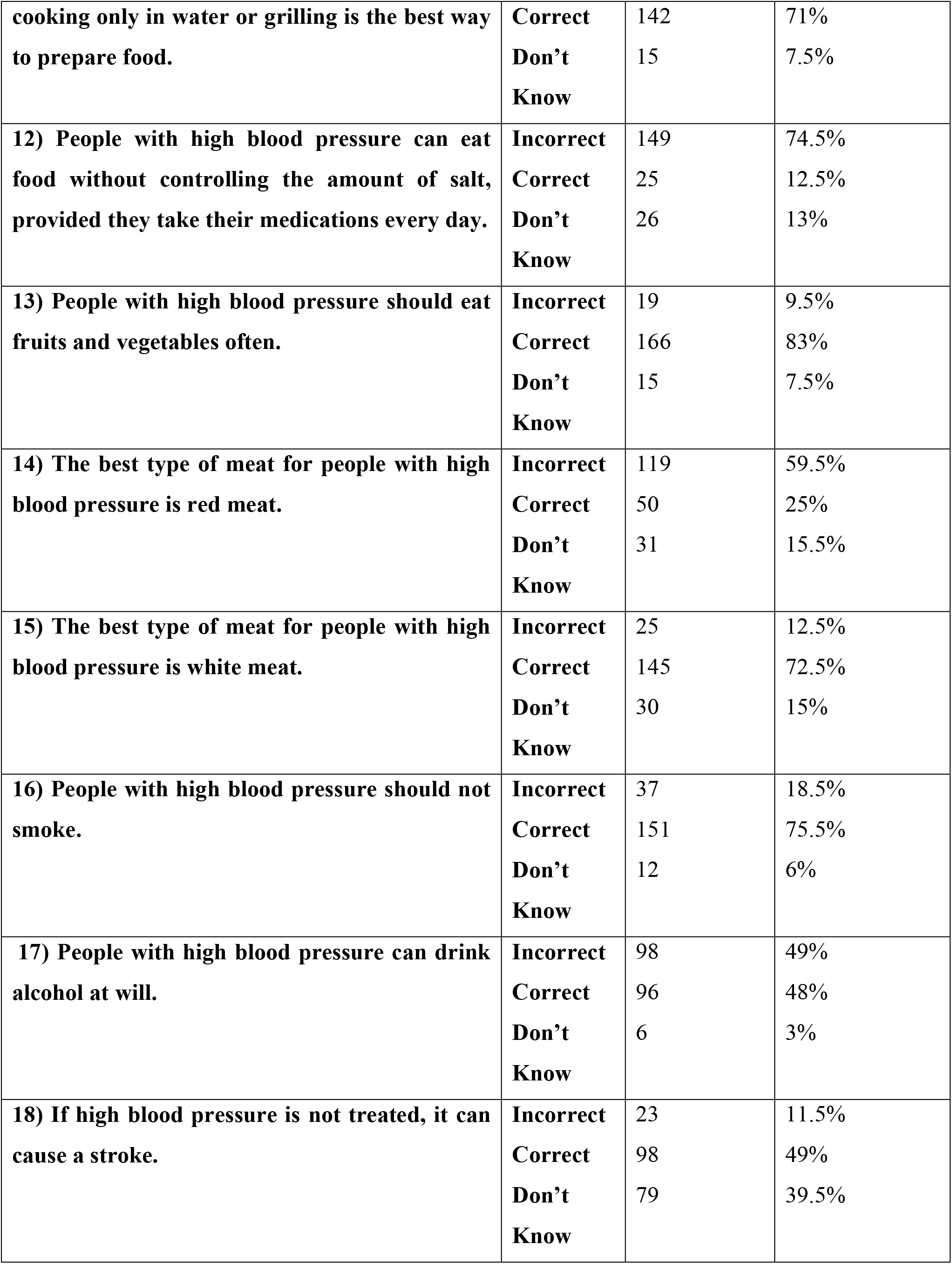
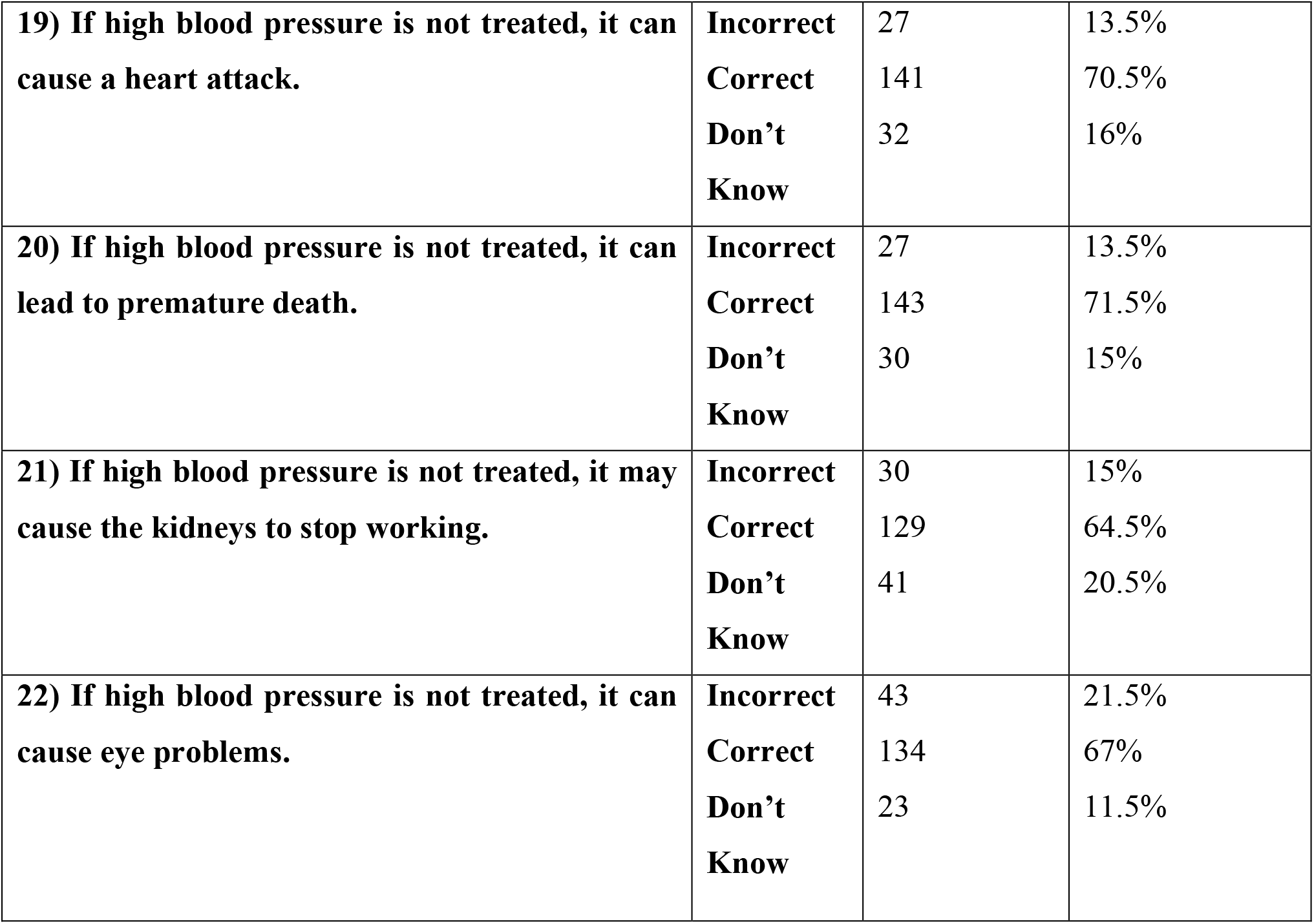
Responses to Hypertension Knowledge Level Scale Components.

A critical misconception was identified in the Drug Compliance sub-dimension, where 67% of participants incorrectly believed that age-related high blood pressure does not require treatment. Similarly, in the Lifestyle sub-dimension, 79% incorrectly identified frying as a healthy cooking method. However, knowledge was strong regarding fruit and vegetable consumption (83% correct) and smoking risks (75.5% correct).

Regarding the Complications sub-dimension, awareness of the risk of heart attack (70.5% correct) and premature death (71.5% correct) was high. However, knowledge regarding the risk of stroke was notably lower, with only 49% of participants providing the correct answer. The overall average number of correct answers was 14.1 out of 22 (64.5%).

As a whole, these results imply that some people are familiar with certain attributes linked with hypertension, but nevertheless, substantial knowledge gaps continue to exist, especially on non-adherence to medical treatment regimens and lifestyle implications, as well as serious potential complications associated with uncontrolled blood pressure. This, therefore, calls for specific educational interventions aimed at addressing such misconceptions, hence improving overall patient management of high blood pressure.

## DISCUSSION

Hypertension (HTN) is a major contributor to the high incidence of cardiovascular diseases (CVD), the leading cause of death globally. Hypertension is expected to account for up to 7.6 million (12.8%) of the total number of fatalities each year [9]. In addition to the established risks of stroke, heart attack, and renal disease, hypertension can contribute to the development of atherosclerosis, which is a key factor in vascular diseases like Subclavian Steal Syndrome (SSS). Understanding these relationships is critical for patients to comprehend the whole range of cardiovascular risks associated with hypertension [10]. This study assessed knowledge of hypertension and related variables among hypertensive patients. Our findings demonstrate varying degrees of understanding, identifying both strengths and crucial knowledge gaps in hypertension care that are consistent with and contribute significantly to current research. Our study found a moderate overall knowledge level (mean score 14.1/22), with specific strengths and weaknesses. While a majority of participants correctly identified high systolic (61%) and diastolic (64.5%) pressures as hypertension, a substantial minority (12%-17.5%) answered incorrectly, paralleling findings from a study in Ethiopia, which also noted gaps in fundamental hypertension knowledge [11]. The finding that a significant portion of the participants recognize the requirement for ongoing treatment (83.5% right in Component 6) is encouraging. However, over half of the participants had misconceptions about medication use during symptomatic times. Researchers found that incorrect assumptions regarding medication use during asymptomatic periods can substantially undermine effective hypertension management [12]. Moreover, the Drug Compliance sub-dimension found that 67% of participants incorrectly believed that hypertension in older adults did not require medication. Similar misconceptions were highlighted, indicating a general underestimate of hypertension’s risks among older persons and emphasizing the significance of targeted educational initiatives for this demographic [13]. A particularly alarming finding was the dietary misconception that frying is a healthy cooking method, held by 79% of our participants. This is significantly higher than rates reported in a similar study in urban India, which found dietary knowledge to be poor, but not this specific misconception, to be so prevalent [14, 15]. This suggests a culturally specific educational need in our population. In contrast, awareness of the benefits of fruits and vegetables (83% correct) and the risks of smoking (75.5% correct) was high, indicating that some public health messages are effective. Only over half of respondents were aware of the risk of stroke, despite our results showing moderate to high awareness of the major sequelae linked to untreated hypertension. A study that highlighted the need for improved patient education on the numerous hazards of untreated hypertension, especially stroke, has previously identified and examined this particular knowledge gap [16]. Accurate evaluation of hypertension knowledge is an important initial step in identifying individuals in need of hypertension education, because knowledge is frequently required for a patient to undertake optimal hypertension self-care, as demonstrated by prior studies [17, 18].

This study has several limitations. First, the cross-sectional design prevents the establishment of causality between knowledge and health behaviors. Second, the use of a non-probability convenience sample from a single tertiary care hospital limits the generalizability of our findings to the broader Pakistani population. Third, the data collection was conducted over a narrow one-month period, which may not capture seasonal or other temporal variations. Finally, as this was a descriptive study, we did not perform multivariate analysis to identify predictors of poor knowledge; this remains a key area for future research. Future studies should employ longitudinal designs and regression models to explore the factors influencing hypertension knowledge and its impact on clinical outcomes over time.

## CONCLUSION

In conclusion, the study reveals varying levels of awareness about hypertension among cardiology patients, but also highlights gaps in patient education. Misconceptions about medication and dietary adjustments persist, emphasizing the need for targeted interventions. Enhancing hypertension literacy can improve patient outcomes and reduce cardiovascular disease burden globally. Healthcare providers should invest in comprehensive patient education programs to empower patients.

## Data Availability

None

## Declarations

### Conflict of Interest /Competing interests

NONE

### Funding

NONE

## Acknowledgments

NONE

## Ethics approval

Taken from the LUMHS Ethical Review Committee (LUMHS/REC/-312).

## Written Consent for publication

Taken through the Consent Form

## Availability of data and material (data transparency)

None

## Code availability (software application or custom code)

None

## Consent to participate

Informed consent was obtained.

## Author’s Contribution

1. **Fatima Khurshid:** Study Concept and Design, Drafting Article, Data Interpretation
2. **Muhammad Owais:** Data Collection, Accountable for all aspects of the work
3. **Iqra Tahir Dar:** Literature Review
4. **Fiza Ameer:** Data Collection
5. **Absar:** Data Collection
6. **Ayesha Khurshid:** Data Analysis
7. **Raheel Ahmed:** Corresponding Author, Final Review
8. **Bravein Amalakuhan:** Supervision

## Notes

### Competing Interest Statement

The authors have declared no competing interest.

### Author Declarations

Taken from the LUMHS Ethical Review Committee (LUMHS/REC/-312).

